# Examining practicality of current procedural terminology standard codes for privacy of patients at Rwanda Military Referral and Teaching Hospital and Legacy Clinics

**DOI:** 10.1101/2024.12.03.24316880

**Authors:** Nemeyimana Patrick, Uwitonze Alfred, Ingabire Eliane, Sugira Vicent, Mugisha Emmy, Ruhumuriza Anselme, Muvunyi Z. Thierry, JMV Gapira Ganza, Usengimana Angelique, Uwihirwe Mickal, Uwumuremyi Fabrice, Mpinganzima Lydivine, Bisanukuri Evergiste

**Affiliations:** University of Rwanda, College of medicine and health sciences,EAC regional center of excellence in biomedical engineering and eHealth,Medical informatics Innovates Ltd,University of Rwanda, College of medicine and health sciences; University of Rwanda, College of medicine and health sciences; Rwanda Military Referral and Teaching Hospital, Research and Innovation; Legacy clinics and diagnostics ltd

**Keywords:** Current procedural terminology, privacy, legacy clinics, Rwanda military hospital

## Abstract

**BACKGROUND:** Emphasizing the critical role of standardized codes in modern healthcare systems, particularly Current Procedural Terminology (CPT) codes, which facilitate efficient communication, accurate billing, and comprehensive patient record management, is of paramount. Despite their benefits, a significant concern regarding patient privacy amidst the detailed documentation enabled by CPT coding. As healthcare providers adopt these standards, they must navigate complex regulatory landscapes like HIPAA to ensure patient confidentiality. However, gaps remain in fully implementing privacy guidelines associated with CPT codes, especially in settings like Rwanda Military Referral and Teaching Hospital and Legacy Clinics, where this study aims to evaluate and improve the practical application of CPT standards to protect patient privacy effectively.

**AIM:** The main objective of this study was to examine a level to which Current procedural terminology standards codes utilized as it can protect privacy and confidentiality.

**METHOD:** Descriptive research design that employed quantitative approaches was used. The research focused on assessing the implementation of Current Procedural Terminology (CPT) standards across two healthcare settings: Rwanda Military Referral and Teaching Hospital and Legacy Clinics. Purposive sampling was employed to select these study sites based on their accreditation and quality service standards relevant to privacy measures and information management. The study population consisted of 177 participants, including medical doctors, insurance partners, Teller staff, and IT officers, selected through stratified and proportionate sampling methods. These methods were chosen to collect quantitative data on the practicality and efficiency of CPT standards in daily healthcare operations. Data management and analysis were conducted using Microsoft Excel for organizing questionnaire responses and STATA software for descriptive analysis.

**Findings:** The findings reveal that the adoption of Current Procedural Terminology (CPT) codes in healthcare facilities is generally low, with Rwanda Military Hospital (RMH) showing no usage and Legacy Clinics having a higher adoption rate of 22.22%. Despite the overall familiarity with CPT, 94.59% of respondents reported non-usage, indicating substantial barriers such as lack of awareness, inadequate training, and systemic challenges. The occupational analysis shows that doctors, who form the largest group, express the highest dissatisfaction rates regarding CPT processes, although the chi-square test reveals no significant relationship between occupation and CPT usage. This suggests that CPT adoption is influenced more by facility-specific issues than by professional roles. Logistic regression analysis highlights that knowledge about CPT negatively impacts its usage, suggesting that awareness alone is not enough to increase adoption. This may be due to insufficient support or practical training. The study suggests that increasing awareness, improving training, and overcoming institutional barriers will be key to enhancing CPT adoption.

**Conclusion:** CPT adoption remains low across surveyed healthcare facilities, with substantial barriers hindering its implementation, particularly at RMH. Occupational roles do not significantly influence usage, but systemic challenges and inadequate training are key obstacles. To promote CPT integration, healthcare institutions need targeted interventions, improved awareness, and comprehensive training programs to ensure successful adoption.

## CHAPTER 1: INTRODUCTION OF THE STUDY

This introduction chapter contains the study background; it also consists of a problem statement, objectives, and questions of the study. Furthermore, it states the significance.

In contemporary healthcare systems, the utilization of standardized codes plays a pivotal role in ensuring efficient communication, accurate billing, and effective management of patient records. Among these standards, Current Procedural Terminology (CPT) codes serve as a universal language facilitating the documentation and billing of medical procedures and services. However, alongside the undeniable benefits of CPT coding, there arises a pressing concern regarding the privacy of patients. As healthcare providers navigate the intricate landscape of medical coding, they must balance the practicality of CPT standards with the imperative to safeguard patient confidentiality.

According to (1), the implementation of CPT codes offers a streamlined approach to recording and reporting medical procedures across diverse healthcare settings. Developed and maintained by the American Medical Association (AMA), CPT codes provide a standardized framework for describing medical, surgical, and diagnostic services. He continued saying that this uniformity enhances the efficiency of healthcare operations, enabling accurate billing, facilitating data analysis, and supporting quality improvement initiatives. Nonetheless, as healthcare organizations adopt CPT coding practices, they encounter challenges related to patient privacy, particularly concerning the sensitive nature of medical information.

As stated recently by (2), that Maintaining patient privacy within the context of CPT coding requires meticulous attention to detail and adherence to established protocols. Healthcare professionals must navigate a complex landscape of regulations, including the Health Insurance Portability and Accountability Act (HIPAA), which sets stringent standards for protecting patients’ medical information. The inherent specificity of CPT codes, while advantageous for coding accuracy, also poses a risk of inadvertently disclosing sensitive details about patients’ health conditions or treatments. He concluded that, consequently, healthcare providers face the ongoing challenge of balancing the need for detailed documentation with the imperative to uphold patient confidentiality.

As technology continues to evolve within the healthcare industry, novel solutions emerge to address the intersection of practicality and privacy in CPT coding(3). Electronic health record (EHR) systems offer sophisticated functionalities designed to enhance coding accuracy while safeguarding patient privacy. Advanced features such as role-based access controls, encryption protocols, and audit trails contribute to the secure handling of sensitive medical data. Furthermore, ongoing advancements in data analytics and artificial intelligence hold promise for refining coding processes, identifying potential privacy breaches, and mitigating risks proactively(4). By harnessing technology responsibly, healthcare providers can navigate the complexities of CPT coding while safeguarding patient privacy in an increasingly digitized healthcare landscape.

In response to these concerns, healthcare stakeholders have called for greater transparency and accountability in the use of CPT codes which seems to be a gap rendering privacy and confidentiality. Organizations such as the American Medical Association (AMA) have developed guidelines and best practices for the ethical use of CPT codes to mitigate privacy risks. These guidelines emphasize the importance of data anonymization, access controls, and encryption techniques to safeguard patient information while ensuring the utility of CPT standards in healthcare operations. As stated by many authors, facilities are under process of implementing the guidelines but few has adored to these protocols.

### 1.1. BACKGROUND OF THE STUDY

Privacy of patients through the use of standards terminologies in healthcare setting has contributed a lot towards medical records(1). The use of the International standard codes for clinical procedures through linking a diagnosis with its codes in every patient journey overcame the bias and brought evidence based diagnosis(2).

Implementation of a new French coding system of clinical procedures, the Classification Commune Des Actes Medicaux (CCAM),along with clinical procedures terminologies which has been developed at the turn of the millennium (between 1996 and2001),it has demonstrated the harmonization and standards of clinical data(3).

Clinical coding has shown importance not only clinically but also on financial implications, and discrepancies in the assigned codes which directly affected the funding of a department and hospital as noticed in the coding of oral and maxillofacial(OMF) procedures(4). Patients ‘information was associated with the use of electronic health records, including privacy, confidentiality, security and patient safety. Given the widespread implementation of electronic health records, there are concerns about data integrity that could jeopardize healthcare quality (5).

According to (5), designing a secure and seamless system that ensures quick sharing of health- care data to improve the privacy of sensitive health-care data, the efficiency of health-care infrastructure, effective treatment given to patients and encourage the development of new health- care technologies. Researchers has shown positive outcome as achieved through the proposed system, a “privacy- aware data tagging system using role-based access control for health-care data(6). Codifying healthcare data into different tags based on International standard codes for specific diagnosis and applying varying levels of anonymization along with role-based access policies is unique to the system.

### 1.2. PROBLEM STATEMENT

Medical records must collect all data concerning in-hospital management of patients: data have to be verified and easily retrievable (8). The introduction of electronic health record systems along with integration of standard terminologies such as current procedure terminologies with added diagnostics standards has contributed to evidence based practice basing on guidelines and privacy of patients data(8). Computerization can offer many advantages to clinicians, but needs some significant adjustments: training and motivation of operators, arrangement of clinical processes and of administrative rules to technological developments.

Nevertheless, some important results can be afforded: standardization of procedures, distribution univocal, verified and ubiquitous data to all concerned operators, protection against undesired retrieve reliability of effective reports (11). In clinical settings, laboratory exams and any clinical procedural that a doctor request, is hand written for billing to be made with cashiers, who is non-medical professional Most of the sensitivepatient information concerning diagnosis especially laboratory exams and clinical procedures must be known by treating personnel; however, they are Viewed by cashiers’ payment who might break the privacy of patients. This study implied-examining the practice coding with current procedure terminologies.

Combination clinical procedures, so that each clinical procedure, diagnosis and laboratory test is coded w standards code for sake of patient’s privacy and confidentiality to unauthorized individual. In Rwanda, study yet was conducted about implementation of current procedural terminology standards codes usability for protecting privacy of patients. This brought my interest to examine practicality of Current procedural terminology standards codes for privacy of patients at Rwanda Military Referral and Teaching Hospital and Legacy clinics as public teaching hospital and legacy clinics as private clinic which have accredited and standardized laboratories to examine standards in terms of privacy.

### 1.3. PURPOSE OF THE STUDY

The purpose of this study was to examine the use of codified clinical procedures and diagnostics tests using current procedure terminology standards codes for privacy and confidentiality of patients at Rwanda military Teaching and Referral hospital and legacy specialty clinics. This purpose was attained with review the use of standard medical terminologies especially diagnosis and procedures and the status as well as the challenges in implementation were examined . The study outcomes were the insights of the level to which practice is current procedure terminology codes for improved patients’ privacy and confidentiality in healthcare settings. After addressing the challenges, solutions will be highlighted and recommended as this will benefit the patients through improved privacy.

### 1.4. OBJECTIVES

#### 1.4.1. MAIN OBJECTIVE

The main objective of this study was to examine a level to which Current procedural terminology standards codes utilized as it can protect privacy and confidentiality.

#### 1.4.2. SPECIFIC OBJECTIVE

1. To know the status of current procedures terminologies usage in clinical setting.
2. To explore how healthcare providers, IT staff and insurance partners perceive t he use of current procedure terminology standards for patients.
3. To determine factors that negatively influences the implementation use of current procedure terminologies on patients’ clinical procedures and diagnosis.

### 1.5. RESEARCH QUESTIONS

1. To what extent are clinical procedures terminology standards used?
2. How important is it to apply current procedural terminologies to different beneficiaries (healthcare providers, ICT officers, insurance companies)?
3. What are the limiting factors in the application of current procedural terminologies in clinical settings?

### 1.6. SIGNIFICANCE OF THE STUDY

The Rwanda Medical Procedure Coding (RMPC) system was developed by Rwanda Ministry of Health to harmonize procedure coding with an international standard. This exercise is a critical foundation for many important reforms within the health sector, including wide spread implementation of electronic medical records systems, interoperability of provider billing systems with health insurance claims management, and costing/tariff setting for stream lining provider payment mechanisms. (21). Though it is in use is electronic health record systems country wide by ministry of health, no study has been done about its linkage with current procedure terminologies for protecting patients ‘data privacy from being overseen by non-medical personal especially in billing process. So, this study revealed these crucial insights which can be used by MoH and others who are responsible in the implementation and strengthening of patient’s privacy and confidentiality by codifying their clinical procedures and laboratory test with i n t e r n a t i o n a l standards.

## CHAPTER2: LITERATURE REVIEW

### 2.1 THEORETICAL LITERATURE

Current Procedural Terminology, commonly known as CPT, is a standardized medical code set that is utilized to report medical, surgical, and diagnostic procedures and Medical services. Developed and maintained by the American Medical Association (AMA),CPT plays a crucial role in facilitating accurate communication, billing, and reimbursement in the healthcare industry(12). This literature review aims to analyze recent research on the practicality of CPT standards codes for patient privacy, both worldwide and specifically in the context of Rwanda CPT codes are alphanumeric and consist of five characters. The codes are organized into three main categories: Category I, Category II, and Category III. Category I codes cover abroad range of procedures and services, including evaluation and management, surgery, radiology, pathology, and more. Category II codes are used for performance measurement and quality improvement, while Category III codes are temporary codes for emerging technologies, services, and procedures(13).

### 2.2. WORLDWIDE PERSPECTIVES ON CPT CODING AND PATIENT PRIVACY

According to (5) conducted a comprehensive analysis of privacy concerns in CPT coding, emphasizing the need for robust privacy measures to safeguard patient information. Their study highlighted the challenges healthcare providers face in balancing the practicality of CPT standards with the imperative to uphold patient confidentiality. Similarly, (6) explored the implications of CPT coding on patient privacy and confidentiality, underscoring the importance of ethical considerations in medical coding practices. (7) Delved into the role of CPT standards in healthcare, focusing on the delicate balance between practicality and privacy. Their research emphasized the necessity of adopting advanced technologies and encryption protocols to mitigate privacy risks associated with CPT coding. (8) Addressed the challenges of ensuring patient privacy in CPT coding, proposing innovative solutions to enhance data security and confidentiality in healthcare operations.

In contemporary healthcare systems, the utilization of standardized codes, such as Current Procedural Terminology (CPT) codes, is fundamental for efficient communication, accurate billing, and effect management of patient records (9).

In Rwanda, the adoption of CPT coding standards has been instrumental in streamlining healthcare processes and improving patient care. However, the practicality of CPT coding must be carefully evaluated in the context of patient privacy. (10) conducted a systematic review on the impact of CPT coding on patient privacy in Rwanda. Their study revealed the need for tailored privacy policies and regulatory frameworks to address the unique challenges faced by healthcare providers in the country.

Recent advancements in technology offer promising solutions for enhancing the practicality of CPT coding while safeguarding patient privacy in Rwanda. For instance, the integration of electronic health record (EHR) systems with robust security features can significantly reduce the risk of privacy breaches associated with CPT coding practices(9). Moreover, capacity-building initiatives and training programs for healthcare professionals are essential to ensure adherence to ethical standards and regulatory guidelines regarding patient privacy in CPT coding.

Analyzing the practicality of Current Procedural Terminology (CPT) standards codes for patient privacy is crucial for ensuring ethical and efficient healthcare delivery worldwide, including in Rwanda. Recent research underscores the importance of adopting advanced technologies and implementing stringent privacy measures to mitigate risks associated with CPT coding practices(11). Moving forward, interdisciplinary collaboration between healthcare providers, policymakers, and technology experts is essential to address the evolving challenges and opportunities in CPT coding while safeguarding patient privacy effectively(4).

### 2.3. EVOLUTION OF CPT

CPT has evolved over the years to adapt to advancements in medical practices, technology, and healthcare delivery. The code set undergoes an annual update process to incorporate new codes, revise existing codes, and ensure relevance in the ever-changing healthcare landscape(12).

For instance, in the 2021 update, the AMA introduced new codes for emerging technologies, telehealth services, and COVID-19-related procedures (AMA, 2021). These updates reflect the commitment of CPT to accurately represent the breadth of medical services provided by healthcare professionals(14).

In Rwanda the Rwanda Medical Procedure Coding (RMPC) system was developed by Rwanda Ministry of Health to harmonize procedure coding with an international standard(13). This exercise is a critical foundation for many important reforms within the health sector, including widespread implementation of electronic medical records systems, interoperability of provider billing systems with health insurance claims management, and costing/tariff setting for streamlining provider payment-mechanisms(13)

### 2.4. UTILIZATION AND SIGNIFICANCE OF CPT CODES

CPT codes are integral to various aspects of the healthcare system, including billing, reimbursement, research, and quality improvement. Health insurance companies use CPT codes to process claims and determine appropriate reimbursement for healthcare services. Furthermore, researchers and policymakers rely on CPT data for analyzing healthcare trends, resource utilization, and outcomes (15).

Accurate utilization of CPT codes ensures that healthcare providers are appropriately compensated for their services, facilitates accurate communication between healthcare professionals and insurers, and contributes to the overall efficiency of the healthcare system. Compliance with CPT coding standards is essential for maintaining the integrity of healthcare data and supporting evidence-based decision-making (16).

### 2.5. CHALLENGES AND FUTURE TRENDS IN CPT

While CPT is widely adopted and recognized, challenges exist in its application. Healthcare professionals often face complexities in selecting the most appropriate code due to the evolving nature of medical procedures and the expansive code set. Additionally, ensuring uniform understanding and implementation of CPT codes across different healthcare settings can be a challenge (17).

Looking to the future, emerging trends such as artificial intelligence (AI) and value-based care may influence the evolution of CPT. Integration of AI tools in coding processes could enhance accuracy and streamline coding workflows. Additionally, as the healthcare industry continues to shift towards value-based care models, CPT may undergo further adaptations to accommodate new payment structures and emphasize quality metrics (18)

Real examples is accurate anesthesiology procedure code data are essential to quality improvement, research, and reimbursement tasks within anesthesiology practices. Advanced data science techniques, including machine learning and natural language processing, offer opportunities to develop classification tools for Current Procedural Terminology codes across anesthesia procedures.

Models were created using a Train/Test dataset including 1,164,343 procedures from 16 academic and private hospitals. Five supervised machine learning models were created to classify anesthesiology Current Procedural Terminology codes, with accuracy defined as first choice classification matching the institutional-assigned code existing in the preoperative database (11).The most important feature in model training was procedure text Through application of machine learning and natural language processing techniques, highly accurate real-time models were created for anesthesiology Current Procedural Terminology code classification. The increased processing speed and a priori targeted accuracy of this classification approach may provide performance optimization and cost reduction for quality improvement, research, and reimbursement tasks to anesthesiology procedure codes.

## CHAPTER3: METHODOLOGY

### 1.1. Introduction

This chapter describes the research methodology which was used. It includes Study area, study design, study population, study sample and sampling strategy, data collection methods and procedures and data analysis.

### 1.2. STUDY DESIGN

This study used descriptive research design in cross sectional method by examining different variables relating to implementation of, questionnaires were used to collected data at current time of attending data collection point of time.

### 1.3. RESEARCH APPROACH

Quantitative method for collecting data using questionnaire were used.

### 1.4. INCLUSION CRITERIA

Medical doctors, Insurance partners, Teller staff and IT officers staff at study sites.

### 1.5. EXCLUSION CRITERIA

Staffs that are not medical staff nor insurance partners nor IT officers or those in selected but refuse to accept consent were excluded from participating in study. These include intern students, cleaners, security guards, and drivers.

### 1.6. SAMPLE SIZE

The calculation of samples size was based total staff included in study at study area. The total at

Rwanda Military Referral and Teaching Hospital are 227 and at legacy clinics are 99.

According Yamane’s formula was used to determine the sample size.

At , Confidence level=95%

- Standard Deviation=50% show
- Margin of error=5%
- The z-score will be 1.96 if the confidence level is 95%

With purposive sampling, in population of 326(99 target staff at legacy clinics and 227 at Rwanda Referral and Teaching hospital, **177 sample included in study.** Byproportion, at legacy clinics, if in 326 in total population of 177 participated, among 99 staff at legacy clinics by proportion; 54 staff selected staff at legacy clinics participated in study. Sub proportion in departments; if 99 among selected staff of legacy clinics ,54 participated, by departments, from 4 IT staff,6 insurance staff,56 doctors and 33 Teller staff, by calculation, 2 IT staff,3 insurance staff,31 doctors and 18 Teller respectively participated in study.

At Rwanda Referral and Teaching Hospital, by proportion, if in 326 staff 177 include in study, from 227 staff (11 IT staff,20 insurance staff,141 doctors, and 55 Teller staff),123 staff include in study. By sub proportion in departments selected purposively, if in 227 only 123 participate in study,5 ICT staff,10 insurance staff,76 doctors,29 Teller staff respectively participated in study.

### 3.6. DATA COLLECTION METHOD AND PROCEDURES

There are some practical issues in data collection such as the credibility in the skills of the researcher and costs in terms of time and money and the choice of suitable method depends mainly on the research questions, the sample and the data sources (19). A selected method should allow a researcher to collect information that will answer research questions.

In this study, purposive sampling was done to select the study site and study population. The study sites were selected based on their accreditation standards in terms of quality services for there are standards for privacy measures information management. ICT departments contribute a lot to implementation of IT related standards, Insurance staff contributes a lot to management and follow up of clinical procedures for reimbursement, doctors play a key to prescription and ordering of most clinical procedures, and Teller staff contribute in accessing and billing as well as management of all clinical procedures.

Particular health facility electronic health record in departments by department, was assessed and see if there are modules for this standard to see how privacy is maintained whether or not the standards of current procedural terminology is in use using questionnaire with pre-set short questions were used to gather quantitative data. Questionnaire proved the perception of CPT and challenges for its implementation.

### 3.7. DATA MANAGEMENT AND ANALYSIS METHOD

Microsoft excel was used to organize data from questionnaire then analyzed quantitatively using STATA software.

### 3.8. ETHICALCONSIDERATION

Before starting data collection, the researcher submitted a research proposal to the University of Rwanda, College of Medicine, and Health Sciences (CMHS) Institutional Review Board (IRB) and ethical clearance with reference number CMHS/IRB/191/2024 was obtained. Before collecting data, authorization to conduct study was granted by study sites upon requesting approval from Rwanda National ethical committee and signing non-disclosure agreement.

As this research study involves human beings and patient’s information, ethical compliance was considered. Thus, it is vital to protect participants’ interests ensuring that participation is voluntary as per the consent form standards. The research undertaken with a scientific integrity and confidentiality obeying the laws of the country and the codes of research ethics (20). Before engaging participants in the research, they were informed about the purpose of the study on an introductory consent form and their role as respondents was clarified. About the clinical procedures and medical records observed at concerned study area, the data remained confidential and researcher was prohibited from taking pictures of patient’s identities as data protection consent was signed by researcher.

### 3.9. STUDY LIMITATION

The study covered samples size of 177 participants from two health facilities due to limited resources otherwise increasing sample size and study area is recommended. CPT standards used new terminologies which was not familiar in daily use, was time consuming in explaining the standards.

## CHAPTER 4: FINDINGS

### 4.1. Descriptive analysis of the findings

This descriptive analysis examines the usage of Current Procedural Terminology (CPT) codes in clinic settings, revealing varied adoption levels, with some facilities fully implementing CPT while others la The study explores healthcare providers’ and insurance partners’ perceptions, showing mixed views on the importance of CPT in enhancing privacy. Additionally, it identifies key barriers to implementation including limited training, resistance to change, and resource constraints, highlighting areas needing improvement to enhance CPT adoption and protect patient confidentiality. the following are the key findings.

**Figure 1:**
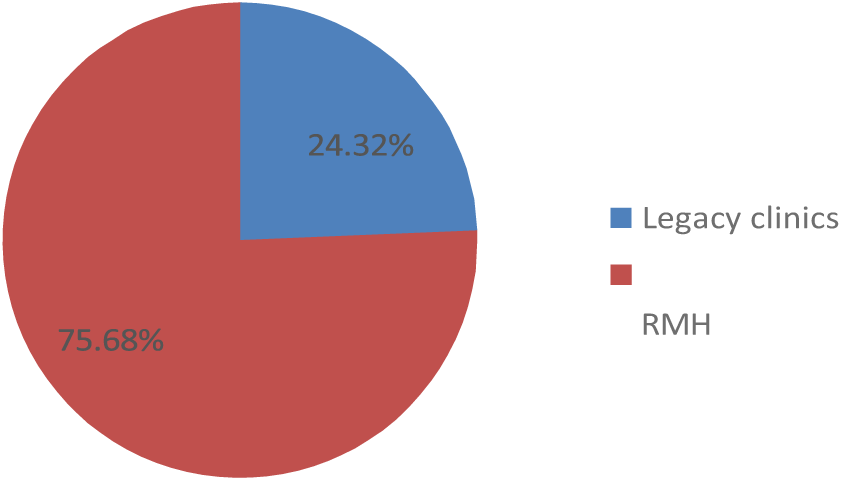
Distribution of Health facility on each category

The distribution of participants across health facilities shows a significant concentration at RMH (Rwanda Military Hospital), with 75.68% of the respondents associated with this facility. In contrast, Legacy Clinics account for only 24.32% of the participants. This distribution indicates a notable imbalance, suggesting that RMH plays a dominant role in the sample, while Legacy Clinics have a relatively small representation. This disparity may reflect differences in facility size, scope, or the types of services offered between the two health facilities.

#### 4.1.1. RESPONSES ON THE USABILITY OF CPT IN HOSPITALS

The diagram above on the usage of CPT (Current Procedural Terminology) indicates that a significant majority of participants, 94.59%, do not use CPT, reflecting a widespread absence of CPT implementation among the respondents. In contrast, only 5.41% of the participants report using CPT, highlighting is limited adoption. This stark disparity suggests that CPT is not commonly utilized in the current practic settings of the majority, pointing to potential areas for intervention or improvement to increase CPT usage.

**Figure 2:**
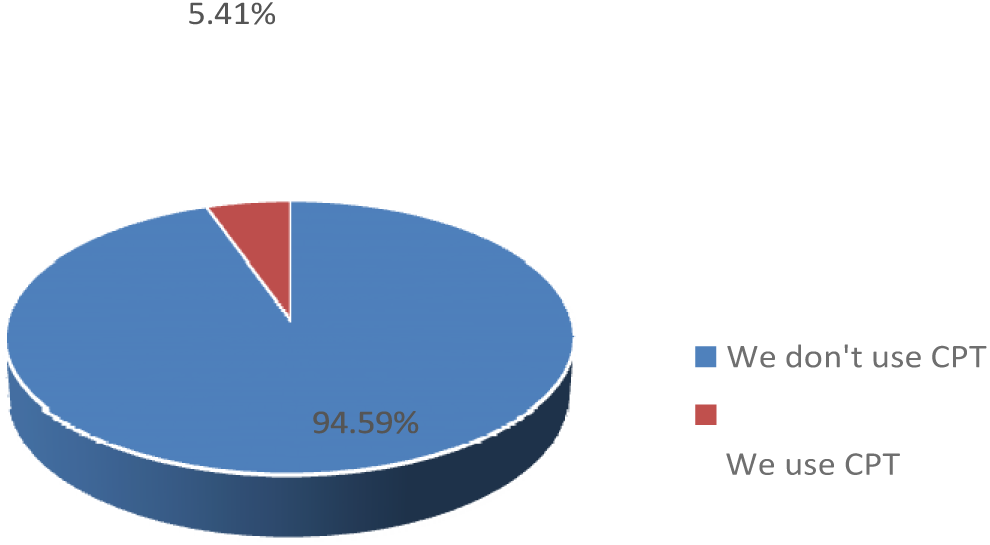
Distribution of CPT usage

The distribution of occupations among the participants reveals a clear concentration in the healthcare an financial sectors. Doctors constitute the largest group, making up 66.89% of the participants, indicating dominant representation of medical professionals in the study. Insurance staff are represented by 10.81% highlighting a significant presence in the sample, though not as large as doctors. The term “insurance appears to be an additional, less common category with only 0.68% representation, possibly indicating more specific subset of the insurance sector. IT staff account for 8.78% of the participants, showing moderate level of representation. Tellers, another occupational group, make up 12.84% of the sample. Th distribution underscores a predominance of doctors and notable representation from insurance-relate roles, with smaller but still relevant participation from IT staff and tellers.

**Figure 3:**
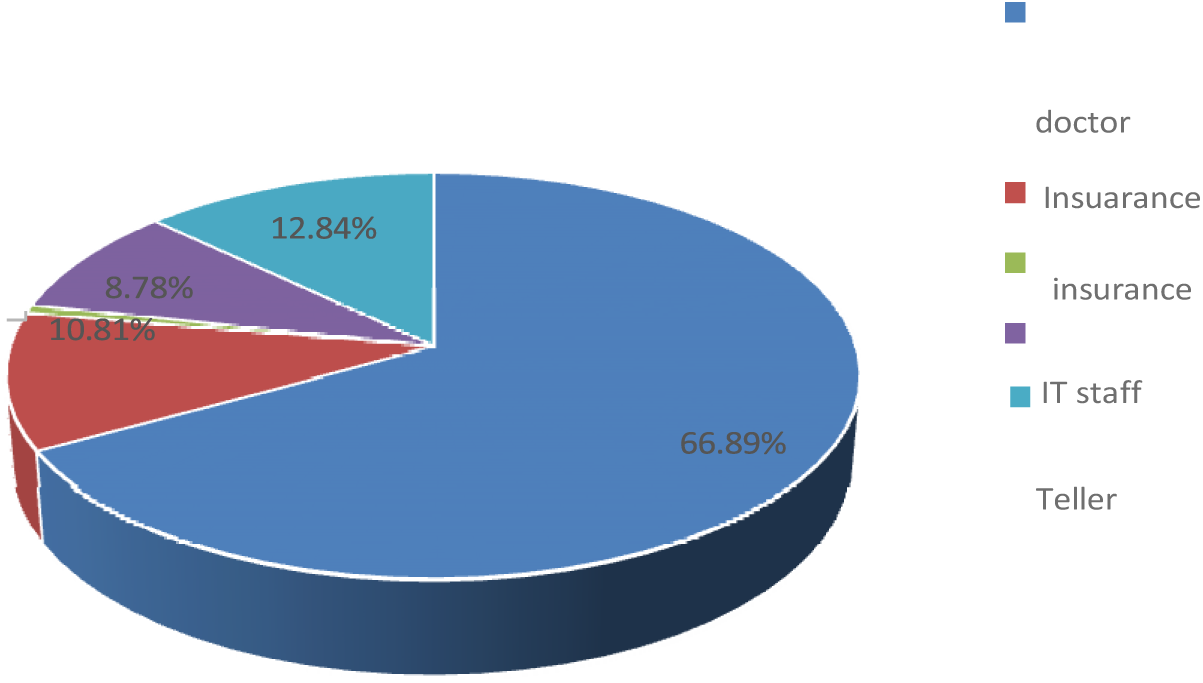
Graph shows % Distribution of occupation

#### 4.1.2. PERCEPTION OF PARTICIPANTS ON THE USE OF SHARABLE PAERS FOR PATIENTS’ DATA

The diagram shows that doctors, making up 66.89% of the occupational group, are the largest segment and 47.30% are employed at RMH (Rwanda Military Hospital). Insurance staff represent 10.81% of the group with a similar dissatisfaction rate of 10.81%. They are less common in Legacy Clinics (1.35%) compare to RMH (9.46%). IT staff account for 8.78% of the participants and also report a dissatisfaction rate 8.78%, with 1.35% in Legacy Clinics and 7.43% in RMH. Tellers constitute 12.84% of the group, an 12.84% express dissatisfaction, with 2.03% working in Legacy Clinics and 10.81% in RMH. The separated category of “insurance” is represented by 0.68%, and their dissatisfaction rate mirrors this lo representation, with a minimal presence in RMH (0.68%). This distribution indicates a notable concentration of dissatisfaction among doctors, insurance staff, IT staff, and tellers, with varying levels representation and dissatisfaction across Legacy Clinics and RMH, suggesting a need for targeted improvements in these professional environments.

**Figure 4:**
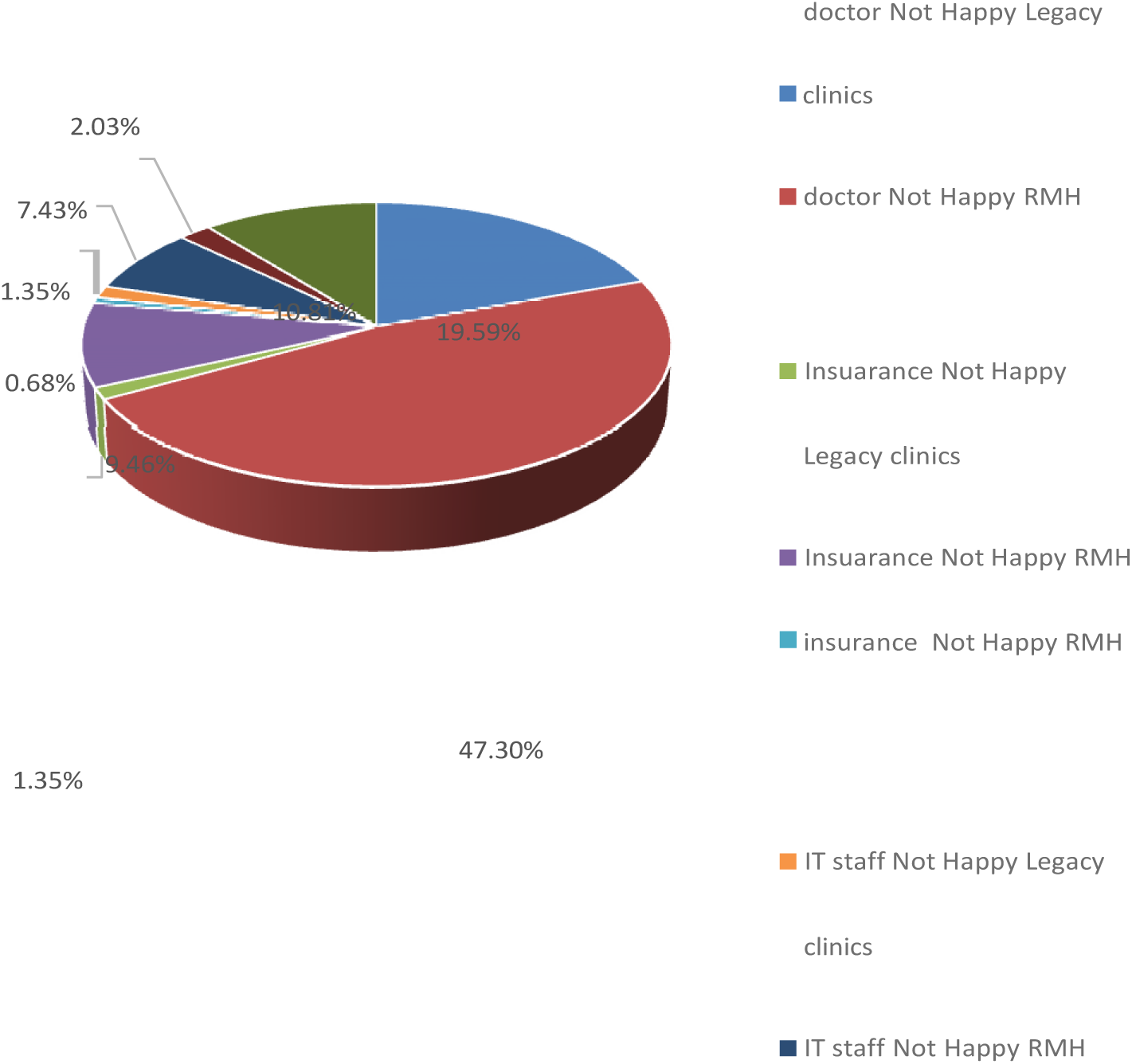
Distribution of occupation, health facilities, with Perceive by recording in paper which is sharable

This graph indicates that a substantial majority of respondents (94.59%) do not use CPT (Curre Procedural Terminology), regardless of their perception of privacy. Among those who are satisfied wit their privacy, only 4.73% use CPT, while 0.68% of those dissatisfied with their privacy report using CP This suggests that CPT usage is very limited overall, and even among those who feel content with the privacy, the adoption of CPT remains low.

**Figure 5:**
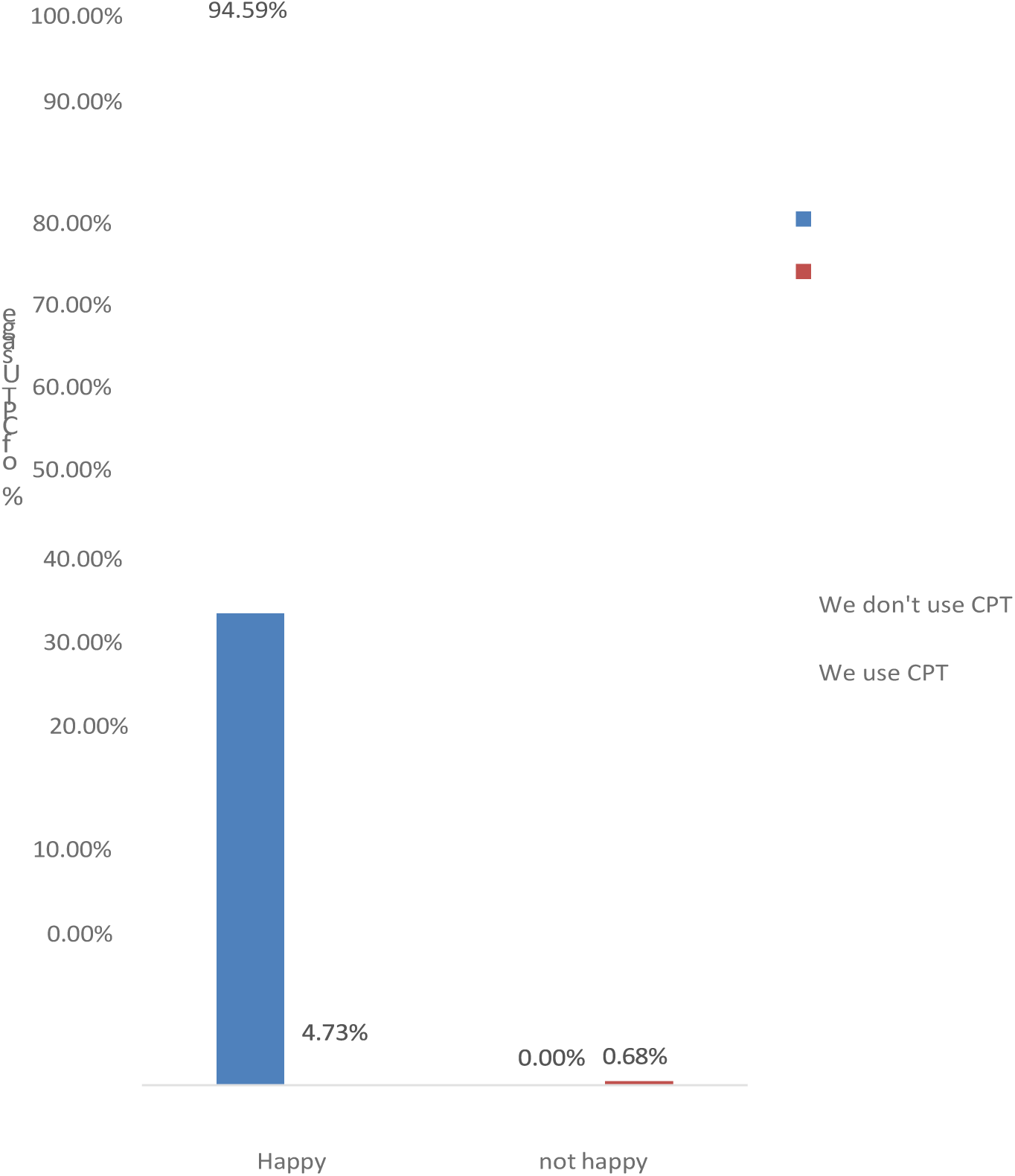
Distribution of perceived treated privacy by CPT usage

**Figure 6:**
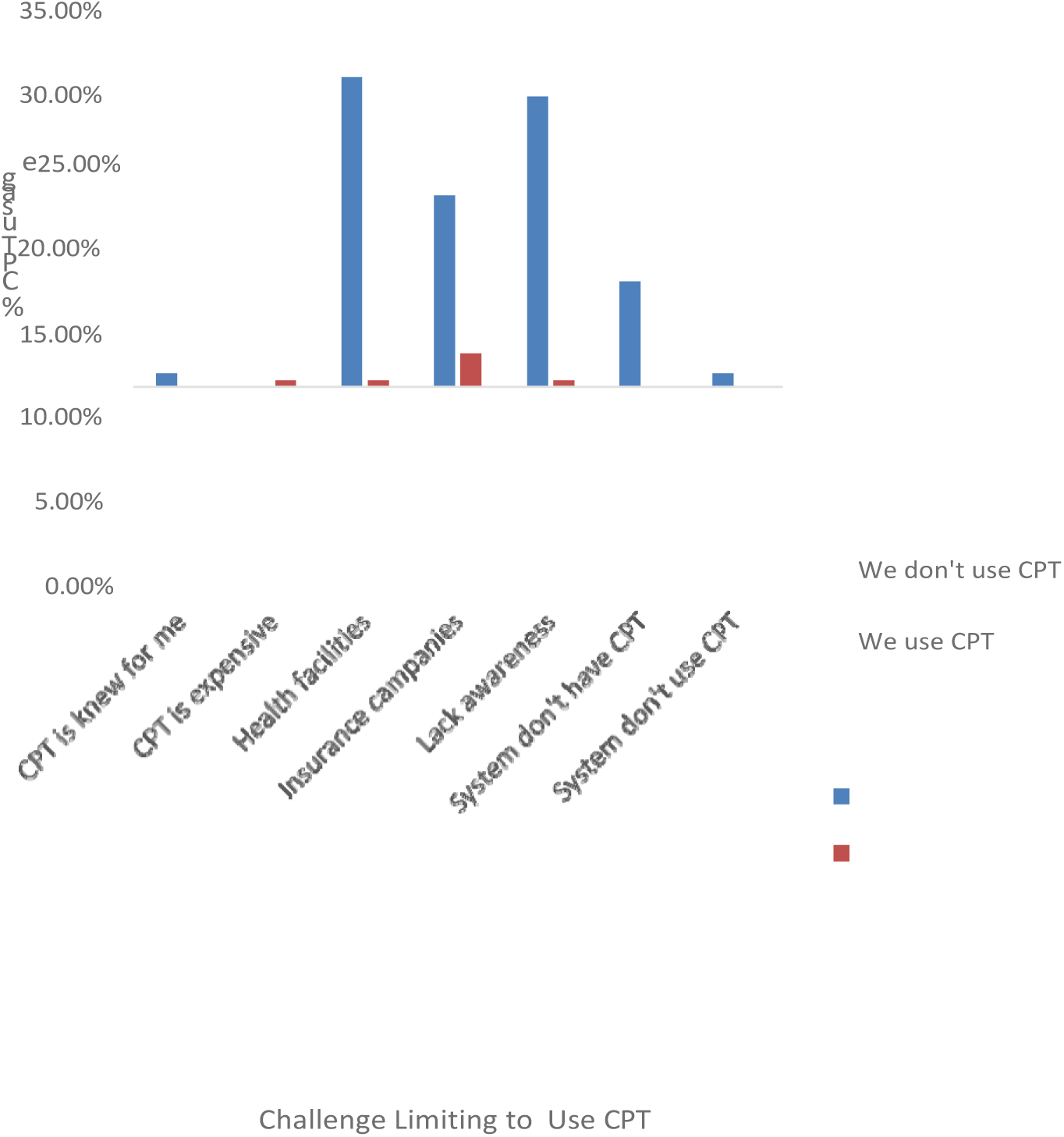
Distribution of CPT usage by challenge limiting to Use of CPT

This graph shows the distribution of responses regarding challenges to the usage of CPT (Curre Procedural Terminology) and reveals that the most significant barriers are related to general issues an institutional factors. Specifically, 29.73% of participants identified a lack of awareness challenge, while 31.76% cited issues related to health facilities. Additionally, 19.59% of as a maj responden pointed to challenges with insurance companies. Only 5.41% of participants use CPT, with a notab proportion of those users encountering issues related to insurance companies (3.38%) and the expense CPT (0.68%). The data also indicates that 10.81% of participants face issues because the system does n majority of respondents (94.59%) do not use CPT, highlighting that awareness and systemic integratio are key areas needing improvement to enhance CPT adoption.

#### 4.1.3. KNOWLEDGE OF HOSPITAL STAFF ON CPT STANDARDS

The Diagram shows that a majority of respondents (58.11%) are unfamiliar with the CPT (Current Procedural Terminology) system, with 57.43% reporting that it is new to them and not in use at the facility. Among those who are aware of the CPT system, 27.03% know about it but have not received and training, with only 2.03% of this group actually using it. Additionally, 14.86% of respondents are familiar with the system, yet only 2.70% of them use it. Overall, CPT usage is quite low (5.41%)

**Figure 7:**
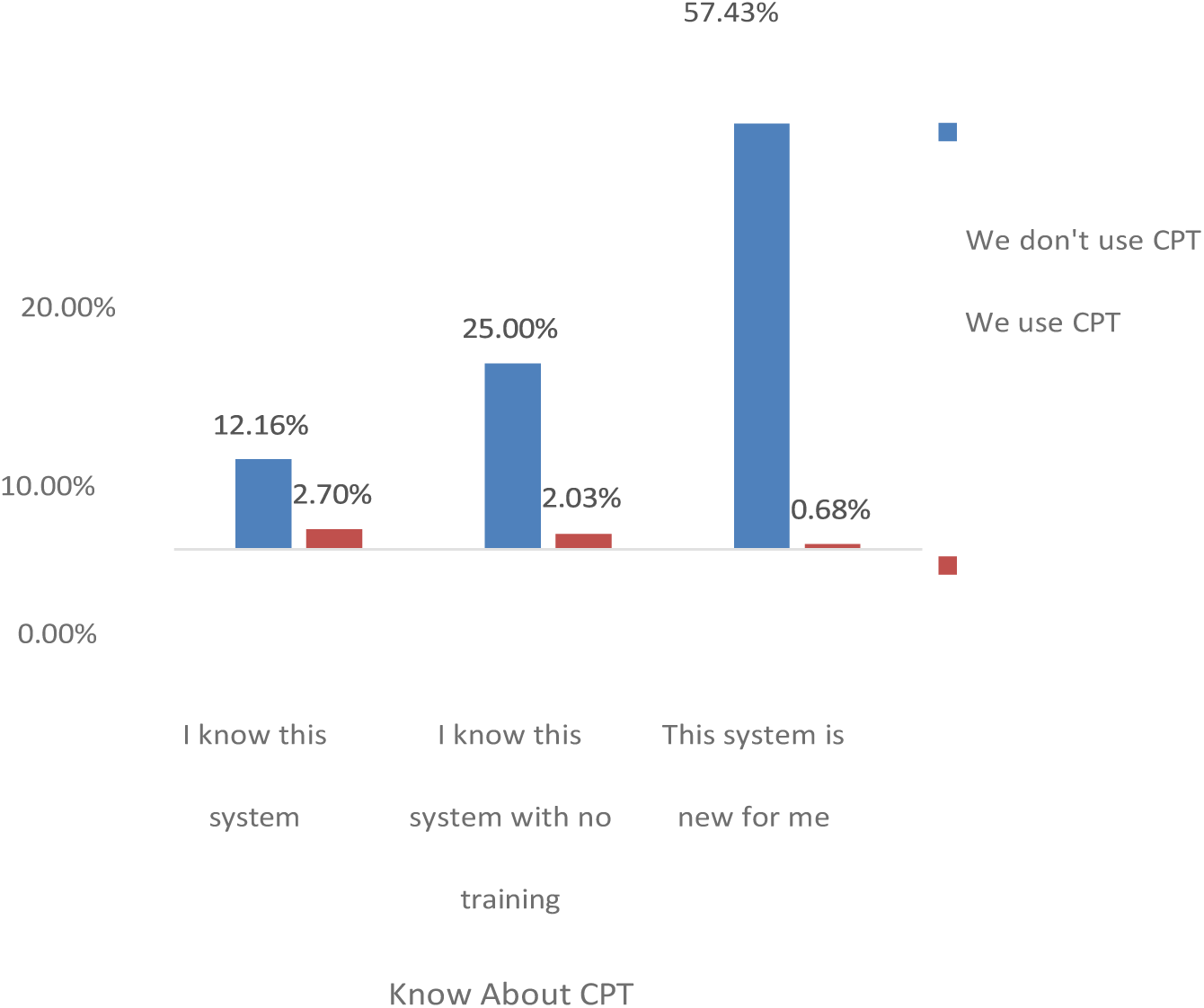
Distribution of CPT usage and Know about CPT

#### 4.1.4. THE CURRENT METHODS USED FOR TREATING PATIENTS’ DATA

Regarding to above graph, Most participants (95.95%) use papers and non-coding systems for treating procedural privacy, while a small percentage (1.35%) use these systems or are unsure of the methods in place. Among CPT users, 50.00% use papers and non-coding systems, while non-CPT use predominantly use these methods (98.57%). Most CPT users (87.50%) are satisfied with their privacy treatment, although 12.50% are not, compared to 100% satisfaction reported by non-CPT users. The CPT adoption if the identified barriers are addressed.

**Figure 8:**
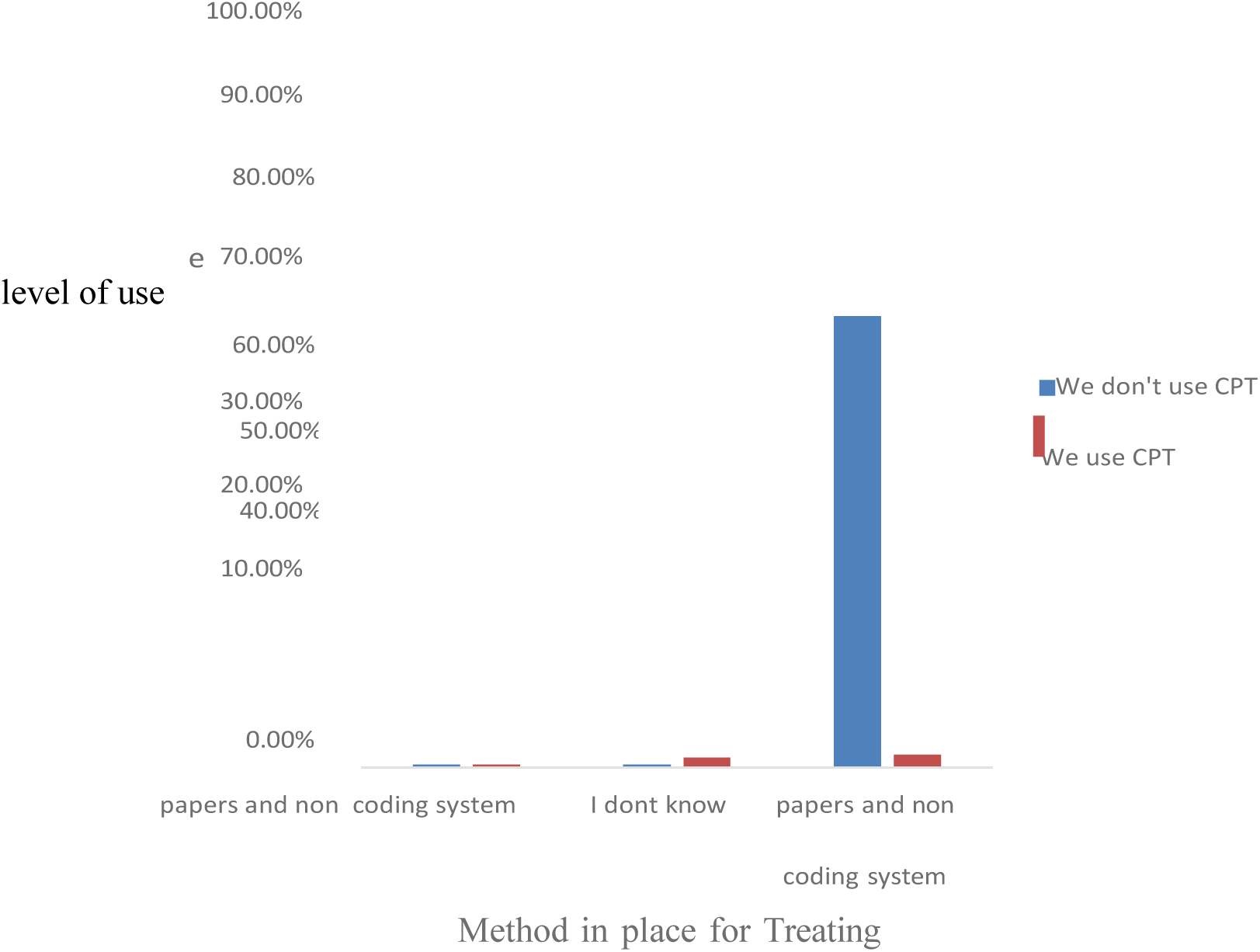
Distribution of CPT usage by Method in place for treating Procedure privacy

**Figure 9:**
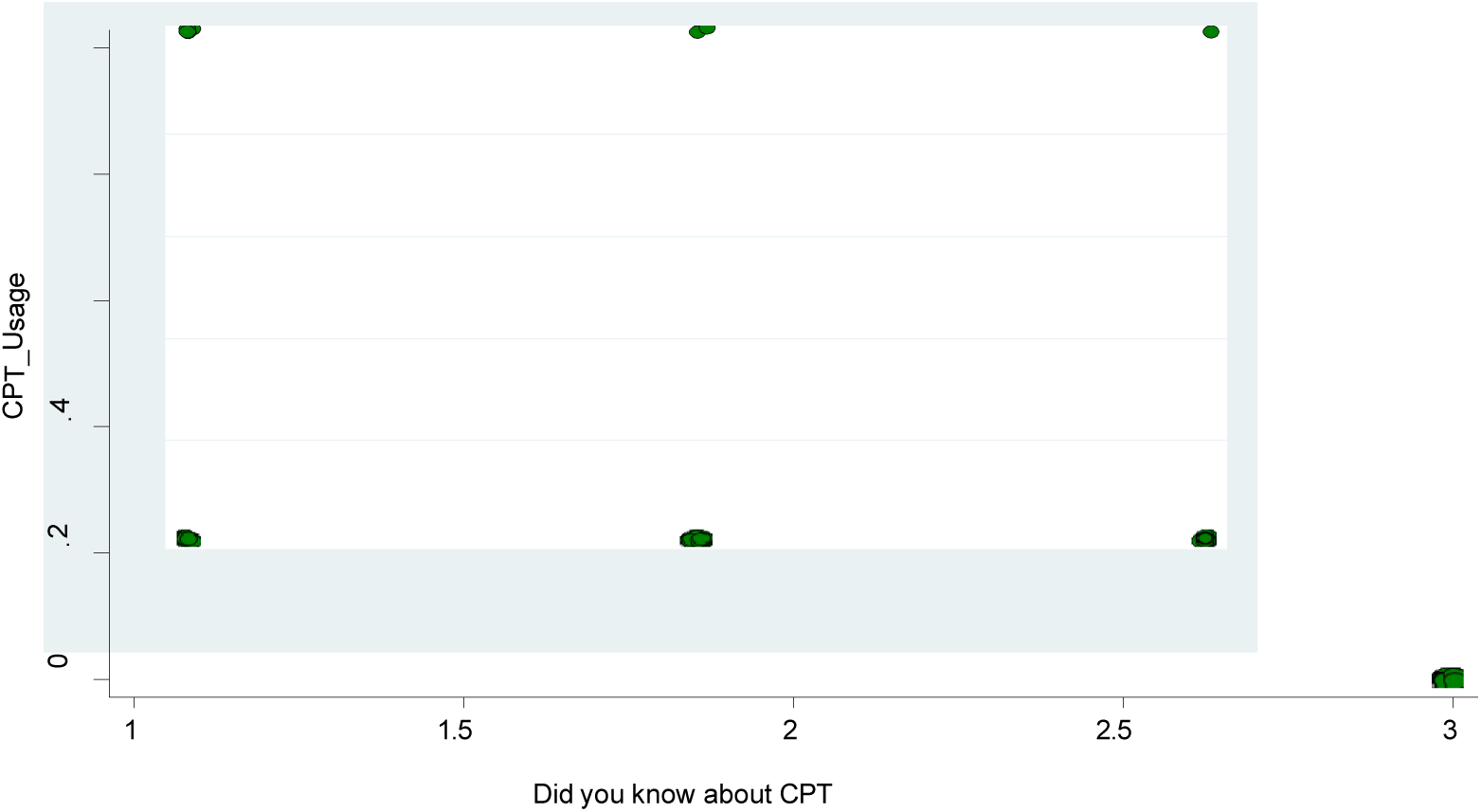
Relationship between knowledge of CPT and the usage of CPT

### 4.2. INFERENTIAL STATISTICS OF THE FINDINGS

**Table 1:**
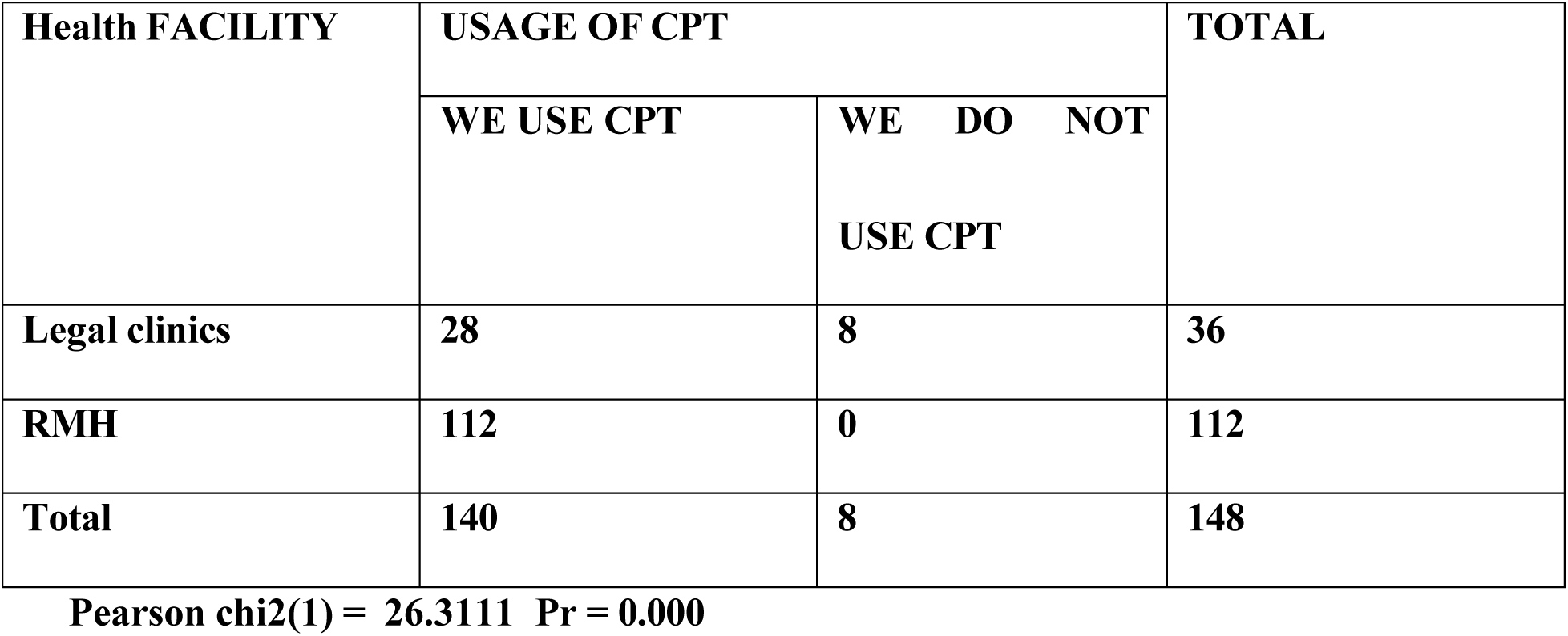
Chi-square analysis.

| Health FACILITY | USAGE OF CPT |  | TOTAL |
| --- | --- | --- | --- |
|  | WE USE CPT | WE DO NOT USE CPT |  |
| Legal clinics | 28 | 8 | 36 |
| RMH | 112 | 0 | 112 |
| Total | 140 | 8 | 148 |

**Pearson chi2(1) = 26.3111 Pr = 0.000**

The chi-square analysis offers significant insights into CPT (Current Procedural Terminology) usage patterns across different health facilities and occupations. For the health facility variable, the chi-squares test produced a Pearson chi-square value of 26.3111 with a p-value of 0.000. This result indicates a highlight significant association between health facility type and CPT usage. Specifically, out of 36 participants from Legacy Clinics, 8 (22.22%) use CPT, while none of the 112 participants from Rwanda Militar Hospital (RMH) use CPT. This disparity highlights that Legacy Clinics have a significantly higher usage rate compared to RMH, suggesting that the type of health facility strongly influences CPT adoption.

**Table 2:** Chi-square test.

| **OCCUPATION** | **USAGE OF CPT** |  | **TOTAL** |
| --- | --- | --- | --- |
|  | **WE USE CPT** | **WE DO NOT**<br>**USE CPT** |  |
| **IT Staff** | **13** | **0** | **13** |
| **Insurance** | **16** | **1** | **17** |
| **Teller.** | **18** | **1** | **19** |
| **Doctor** | **93** | **6** | **99** |
| **Total** | **140** | **8** | **148** |

**Pearson chi2(4) = 0.9062 Pr = 0.924**

In contrast, the chi-square test for occupation and CPT usage yielded a Pearson chi-square value of 0.906 with a p-value of 0.924. This result indicates no significant relationship between occupation and CP usage. For example, out of 99 doctors, 6 (6.06%) use CPT, compared to 1 (6.25%) of 16 insurance staff, (5.26%) of 19 tellers, and no IT staff using CPT. The lack of significant association suggests the differences in CPT usage across occupations are not statistically meaningful, implying that occupation does not significantly affect whether individuals use CPT.

**Table 3:** Logistic regression.

| CPT USAGE | COEFFICIENT | Z_STAISTICS | P VALUE | [95% CONF.INTERVAL] |
| --- | --- | --- | --- | --- |
| The challenge<br>in the usage of<br>CPT | **-.4573399** | **-1.48** | **0.140** | **-1.064869 .1501888** |
| Know about<br>CPT | **-1.488862** | **2.92** | **0.004** | **-2.48836 -.489365** |
| Constant | **2.509574** | **1.28** | **0.200** | **-1.325521 6.344668** |

The logistic regression analysis in the above table indicates that knowledge about CPT has a significant negative effect on CPT usage with a coefficient of -1.4889 (p = 0.004), suggesting that knowledge abo CPT is associated with a lower likelihood of using it. This indicates that awareness alone may not lead t increased usage, potentially due to insufficient support or training. Conversely, ‘challenge in use of CP has a coefficient of -0.4573 (p = 0.140), which is not statistically significant, implying that the challeng reported do not have a strong or significant impact on CPT usage. The model overall is statisticall significant (LR chi-square = 11.09, p = 0.0039) with a moderate fit (Pseudo R² = 0.1782), suggesting th the predictors included in the model have a meaningful, though not overwhelmingly strong, influence o CPT usage.

The scatter plot illustrates the relationship between knowledge of CPT (represented by Know about CP on the x-axis) and the usage of CPT (represented by CPT Usage on the y-axis). The green dots indica that respondents knowledgeable about CPT (Know about CPT = 1) are more likely to use it Knowin about CPT= 1), as seen by the concentration of points at the top left of the plot. In contrast, those who ar less knowledgeable or unsure about CPT (with Know about CPT = 2 ) tend to have lower usage rate (Know about CPT = 0), as reflected by the points clustered at the bottom of the plot. This suggests positive association between knowledge and usage of CPT, where increased awareness likely contribute to higher adoption.

## CHAPTER 5: DISCUSSION OF FINDINGS

The descriptive analysis reveals significant variations in the usage of Current Procedural Terminologies (CPT) codes across different health facilities and occupations. The majority of participants are associate with Rwanda Military Hospital (RMH), which shows no CPT adoption, while Legacy Clinics demonstra a higher adoption rate of 22.22%. This disparity suggests that the type of health facility plays a crucial rol in CPT usage. The overall adoption of CPT remains very low, with 94.59% of respondents reporting no use, indicating that CPT is not widely integrated into current practice settings.

The occupational distribution shows that doctors constitute the largest group, and they also exhibit the highest dissatisfaction rates with CPT-related processes. However, the chi-square test for occupation groups reveals no significant relationship between occupation and CPT usage, suggesting that factors influencing CPT adoption are not strongly related to professional roles. The analysis also highlights the despite some familiarity with CPT, the overall usage remains minimal, with significant barriers such a lack of awareness, institutional issues, and inadequate training.

Logistic regression analysis further emphasizes that knowledge about CPT negatively affects its usage which may be due to insufficient support or training rather than a lack of awareness. The data underscore the need for targeted interventions, particularly at RMH, and improved training programs to addresses systemic challenges. Enhancing CPT adoption requires a comprehensive approach that includes increasing awareness, providing training, and addressing institutional barriers to effectively protect patient confidentiality and improve clinical practices.

## CHAPTER6: CONCLUSION AND RECOMMENDATION

### 6.1. CONCLUSION

The analysis shows that Current Procedural Terminology (CPT) usage is notably low across the surveyed health facilities, with Legacy Clinics demonstrating significantly higher adoption compared to Rwanda Military Hospital (RMH), where no CPT usage is reported. Despite some familiarity with CPT among healthcare professionals, the overall adoption rate remains minimal, with 94.59% of respondents utilizing CPT. This indicates substantial barriers to CPT implementation, including lack of awareness insufficient training, and institutional challenges.

The findings also highlight that occupational roles do not significantly influence CPT usage, suggesting that factors beyond professional backgrounds, such as facility-specific issues and systemic integration play a more critical role. Logistic regression analysis shows that knowledge about CPT, rather than promoting usage, is associated with lower adoption rates, potentially due to inadequate support and training. Addressing these barriers requires targeted interventions to improve CPT adoption. This include enhancing awareness, providing comprehensive training, and addressing institutional issues, particularly RMH. By focusing on these areas, healthcare facilities can better integrate CPT into their practice ultimately improving patient confidentiality and overall clinical efficiency.

### 6.2. RECOMMENDATION

To enhance the adoption and effective utilization of Current Procedural Terminology (CPT) codes, it crucial to focus on increasing awareness and training for healthcare professionals and insurance staff Comprehensive training programs should be developed to educate users on the benefits an implementation of CPT codes. These programs should include practical demonstrations, case studies, an ongoing support to ensure that CPT codes are effectively integrated into clinical practices. Emphasizing the importance of CPT in improving privacy and procedural efficiency will help motivate healthcare professionals to adopt these codes.

Addressing institutional barriers is another key recommendation. Many facilities face systemic challenge that hinder the implementation of CPT codes. Upgrading healthcare facilities’ systems to support CP allocating necessary resources, and ensuring CPT codes are incorporated into existing workflows can help overcome these barriers. Engaging with healthcare leaders to advocate for CPT adoption and addressing any resistance to change will be essential in facilitating a smoother transition.

Finally, targeted interventions at facilities with low CPT usage, such as Rwanda Military Hospital (RMH) should be prioritized. Conducting facility-specific assessments will help identify unique challenges a tailor intervention accordingly. Personalized training sessions, technical support, and resource allocation should be provided to these facilities to address their specific needs. Regular monitoring and feedback will be necessary to ensure successful CPT integration and to improve overall adoption rates. By focusing o these areas, healthcare facilities can better integrate CPT codes, thereby enhancing patient confidentiality and procedural efficiency.

## Data Availability

all available on request

